# A global meta-analysis of effects of green infrastructure on COVID-19 infection and mortality rates

**DOI:** 10.1101/2023.05.08.23289653

**Authors:** Bopaki Phogole, Kowiyou Yessoufou

## Abstract

Evidence of the benefits of greenspaces or greenness to human wellbeing in the context of COVID-19 is fragmented and sometimes contradictory. This calls for a meta-analysis of existing studies to clarify the matter. Here, we identified 621 studies across the world, which were then filtered down to 13 relevant studies covering Africa, Asia, Europe, and USA. These studies were meta-analysed, with the impacts of greenspaces on COVID-19 infection rate quantified using regression estimates whereas impacts on mortality was measured using mortality rate ratios. We found evidence of significant negative correlations between greenness and both COVID-19 infection and mortality rates. We further found that the impacts on COVID-19 infection and mortality are moderated by year of publication, greenness metrics, sample size, health and political covariates. This clarification has far-reaching implications on policy development towards the establishment and management of green infrastructure for the benefits of human wellbeing.

## Introduction

Global human population is changing rapidly following an exponential growth path, putting tremendous pressures on natural resources. Currently, it is approximately 7.9 billion people^1^ and is predicted to reach above 9 billion by 2050 or 11 billion by 2100^2^. In response, nature fights back in various ways to bring down global population to a sustainable level. One of these ways is through global pandemics, e.g., COVID-19. Indeed, the world has been witnessing COVID-19 pandemic since 2020, with over half a million of infection cases and over 20,000 deaths in 2020^3^. In 2022, these figures grew tremendously, reaching over 600 million cumulative cases with over 6 million cumulative deaths^4^. Subsequently, various studies, using different metrics of greenness (the total amount of vegetation in an area), were conducted across the globe to investigate whether greenness act as buffer infrastructure against the spread of COVID-19 infection rates and severity.

However, the findings reported in these studies are mixed^5,6^. For example, ref.^7^ found that a 0.1 increase in NDVI is linked to 4.1% reduction in COVID-19 incidence rate ratio in the USA. A similar pattern was observed using street-level indicators of greenness^8^. The mitigating effects of greenness have also been reported elsewhere: in China and India, an increase in greenness shows a strong negative association with the spread of COVID-19 infections and mortalities^9,10^. These negative effects may be interpreted as follows: activities of the Natural Killer (NK) cells in human body are boosted with frequent exposure to vegetation^3,11^ – NK cells, as part of the immune system, attack to eliminate virus-infected cells^12^. Also, by safeguarding against air pollution, vegetation contributes to lower health risks that may aggravate the severity of COVID-19 infection^13,14^. Additionally, green infrastructure often provides spacious environment for physical exercise, recreation, and social events with reduced chances of person-to-person contact^15,16^. As opposed to these negative correlations between greenness and COVID-19 infection rates, reports of positive correlations are also documented. For example, ref.^6^ found that urban greenspaces were associated with an increase in the spread of COVID-19 infections (see also ref.^15,16^).

These mixed findings could be linked to the differences in how COVID-19 severity was measured, e.g., as hospitalization rates, mortality rates, admission rate to ICU, etc. Additional sources of differences in findings may be linked to differences in sample size, type and number of covariates considered, and choice of statistical tests^17,18^. Furthermore, the mixed findings may be linked to differences in how greenness was measured in different studies. Indeed, greenness was variously measured as street trees, botanical gardens, natural forests and grasslands, and residential gardens or as amount of greenness captured in NDVI or EVI or as quality of green infrastructure^7,8,19,20^. For example, ref.^15^ measured greenness as ‘green space density’ which is the proportion of specific vegetation types in a given spatial unit which they correlated with COVI-19 infection risk measured as ‘venue density’ (number of buildings visited by confirmed COVID-19 positive cases). Since greenspaces are attraction sites, they attract increasing number of visitors, thus increasing the infection risks, and leading to a positive correlation between greenness and infection rate^15^. Furthermore, the mixed findings may be linked to the use of various confounding factors in the model of COVID-19 infection and mortality rates. These factors may be age^21^, ethnicity^22^, and poverty level^23^, among others.

The emergence of conflicting findings presents a challenge with regards to the generalization of the benefits of greenness to human wellbeing in the context of COVID-19 pandemic. In such context, a meta-analysis of existing evidence presents an opportunity to integrate the conflicting reported effects of greenness on COVID-19 infection rates and severity to investigate whether generalization is possible. Scientifically rigorous methodologies are increasingly adopted in various studies to improve the validity of findings and lower between-study heterogeneity. These include the use of larger sample sizes, use of multiple predictors, choice of relevant statistical tests and covariates, and use of fine spatial scales^23-26^. Regardless of these advances, consolidation of measured effect sizes and determination of between-study heterogeneity is still needed. To date, several studies have investigated the relationships between the provision and quantity of greenness and its effects on the spread and severity of COVID-19^5,7,10,20^. However, in the context of conflictual findings reported, a meta-analysis imposes itself or becomes an obligation if we are to clarify how greenness or green infrastructure relates to COVID-19. In the present study, our main objective is to provide such clarifications.

## Results

### Characteristics of studies included in the meta-analysis

A total of 621 studies across the world (Figure 1A) were identified through the search of Scopus, PubMed, and Google scholar platforms. After removing irrelevant and duplicate studies, 25 studies remained, covering Africa, Asia, Europe, and USA (Figure 1B). A review of the 25 full-text articles resulted in a removal of 12 studies that were either review/commentary in nature or did not report the required statistical parameters for meta-analysis.

**Figure 1.**
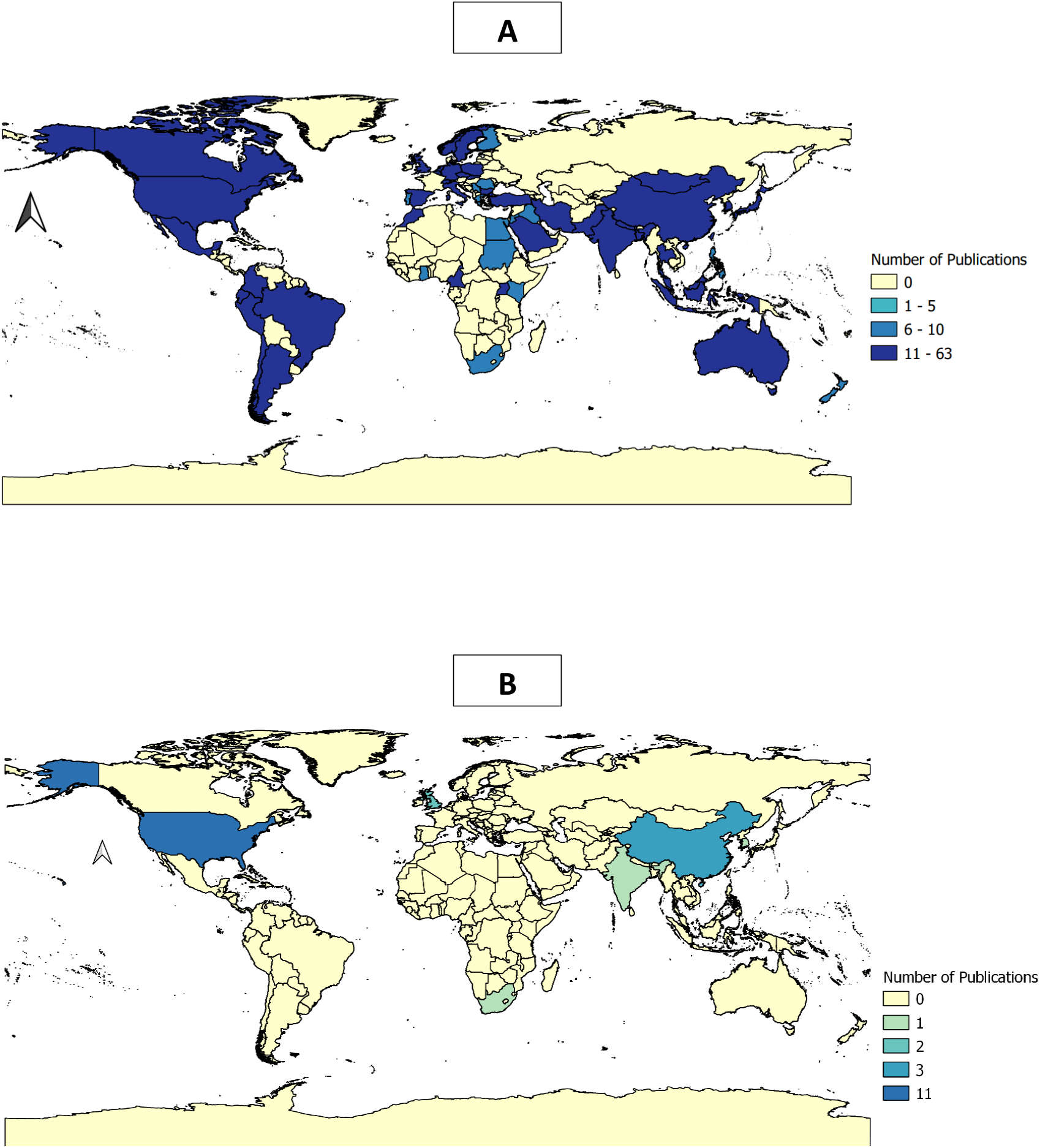
Geography of studies investigating the effects of greenness on COVID-19 infection and mortality rates. (A) Geographical distribution of 621 studies that were retrieved through the search of Scopus, PubMed, and Google scholar; (B) Geographical distribution of studies from our search that focus specifically on the effects of greenness on COVID-19 infections and severity.

The study characteristics are summarized in Table S1. Nine studies that tested the relationships between greenness and COVID-19 infections and four studies that investigated the relationships between greenness and COVID-19 mortality rates were included in the final synthesis. Most of the studies (nine out of 13) were conducted in the United States of America (USA) whereas China, England, India, and South Africa each had one study (Figure 1B). A total of 7 out of 13 studies used more than one predictor of COVID-19 impact in each study with normalised difference vegetation index (NDVI) and abundance of greenness as the mostly used measures of greenness (Figure 2A). Because multiple predictors are used in a single study, a total of 45 different correlations between infection rates and greenness were produced in all 13 studies and 14 correlations between mortality rates and greenness were produced from four studies of COVID-19. We classified covariates into five broad groups: climatic, demographic, economic, health, and political. All 13 studies considered at least one demographic covariate in their analyses, and only four studies included climatic, demographic, economic, health, and political covariates (Figure 2B).

**Figure 2.**
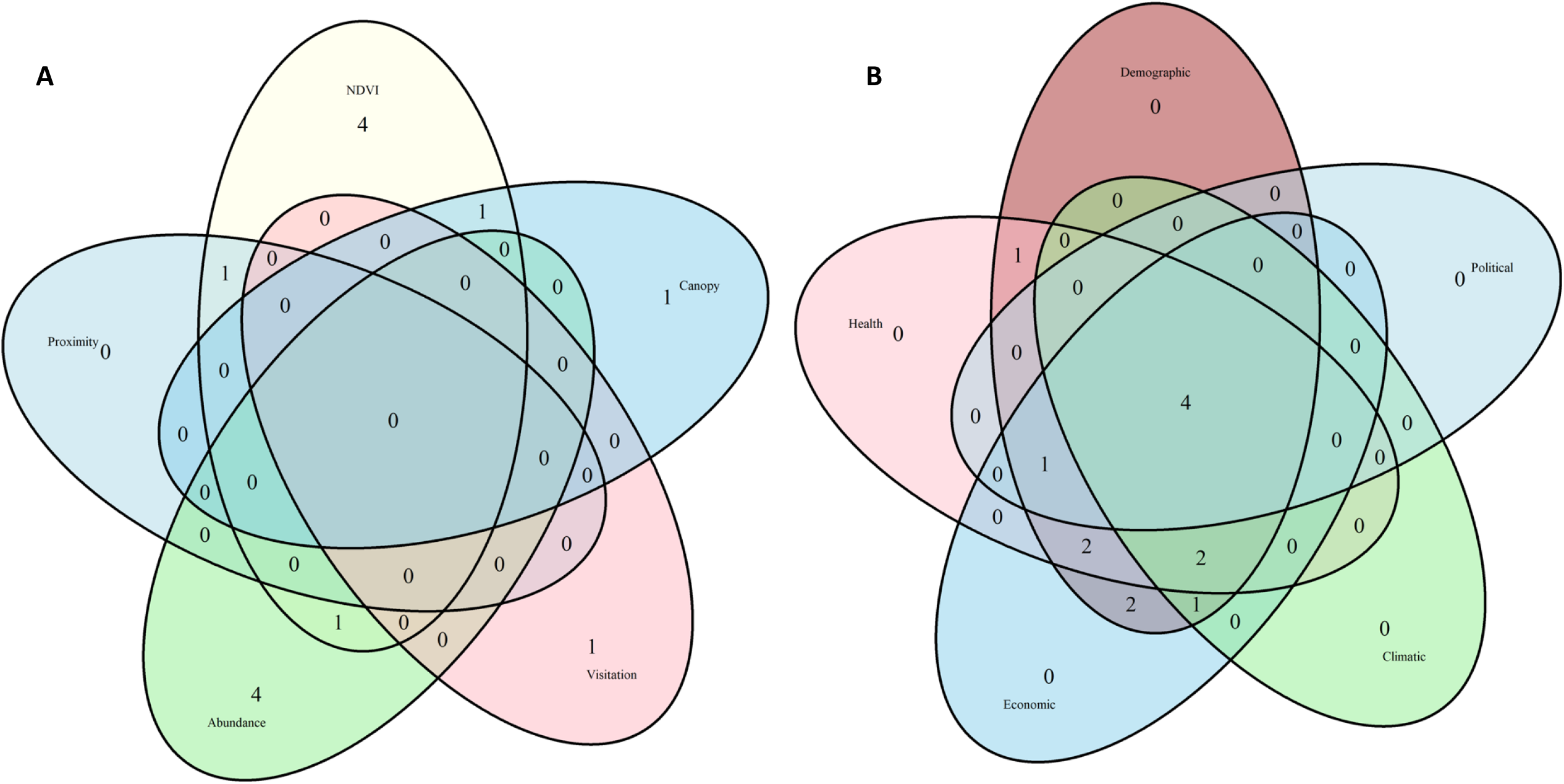
Venn diagram showing the shared factors used in multiple studies that investigate the effects of greenness on COVID-19 infection and mortality rates. A) different metrics of greenness; B) socio-environmental and economic co-variates used those studies.

### Greenness and COVID-19 infections

We found a statistically significant negative effect of greenness on COVID-19 infections (β = −0.08, 95% CI: −0.1396 – −0.0252; t=-2.90; p=0.006) with a prediction interval of [−0.3601 – 0.1954] (95% CI) (Figure 3). Between-study heterogeneity variance was estimated at τ^2^= 0.0184 (95% CI: 0.0185 – −0.0813), with an I^2^ value of 94.1% (95% CI: 92.9% – 95.1%). Subgroup analyses reveal that between-study heterogeneity can be attributed to year of publication (*X*^2^=8.24; p=0.02), choice of predictors (*X*^2^=129.68; p<0.01), and use of political covariates (*X*^2^=8.27; p<0.01) (see Table 1 and Figures S1 – S6).

**Figure 3.**
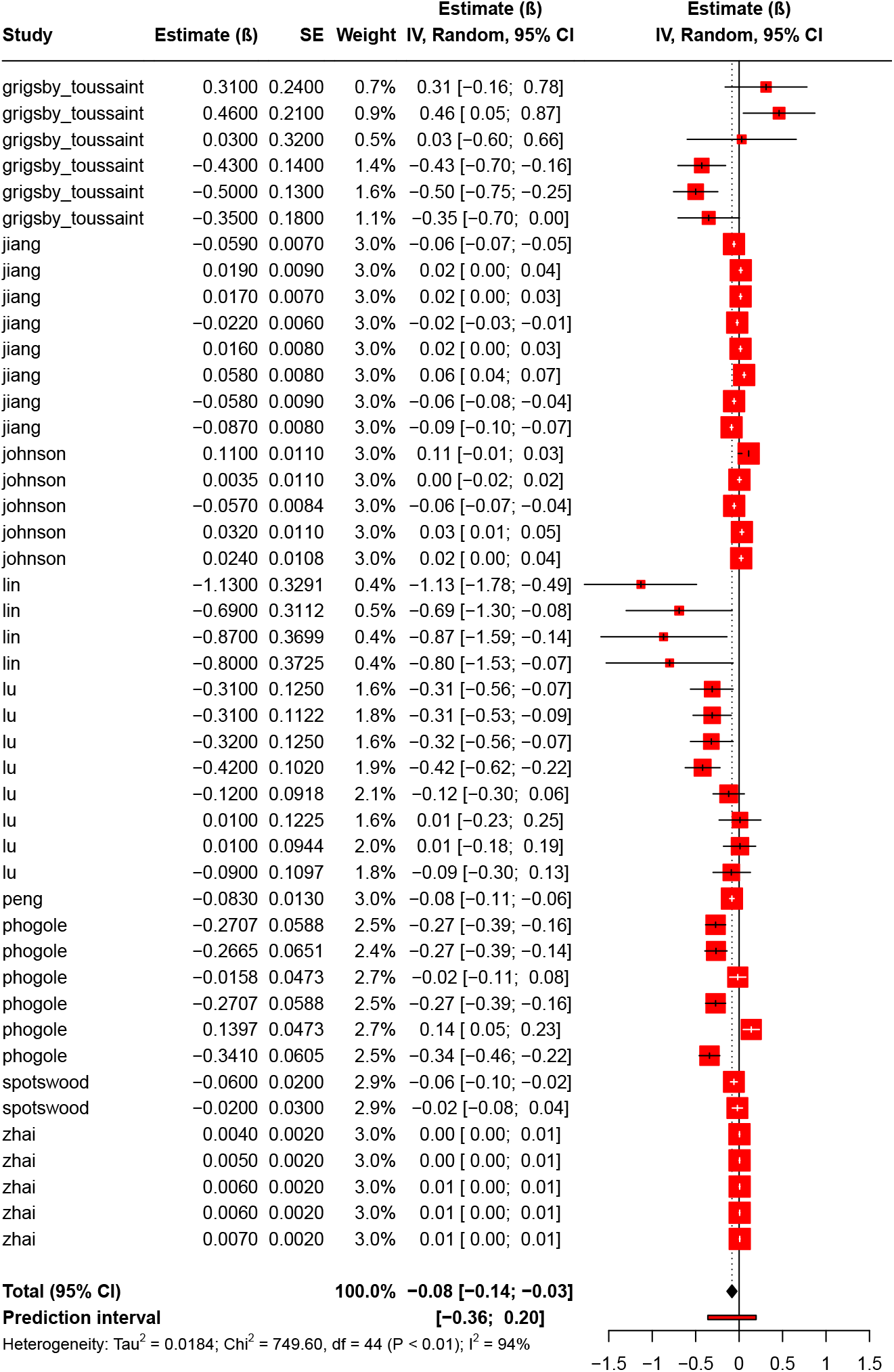
Forest plot of the relationship between greenness and COVID-19 infections.

**Table 1:** Stratified analyses of pooled estimate of COVID-19 infections and green infrastructure.

| Stratified analysis | Number of results | Pooled estimate [95% CI] | Subgroup difference $\chi^2$, df (p-value) |
| --- | --- | --- | --- |
| **Study year** | **45** | **-0.08 [-0.14; -0.03]** | **8.24, df = 2 (p = 0.02)** |
| 2021 | 15 | -0.07 [-0.15; 0.01] |  |
| 2022 | 20 | -0.02 [-0.06; 0.02] |  |
| 2023 | 10 | -0.32 [-0.57; -0.07] |  |
| **Sample size** | **45** | **-0.08 [-0.14; -0.03]** | **0.01, df = 1 (p = 0.91)** |
| Small (n<2000) | 19 | -0.09 [-0.16; -0.02] |  |
| Large (n≥2000) | 26 | -0.08 [-0.18; 0.02] |  |
| **Predictor** | **45** | **-0.08 [-0.14; -0.03]** | **129.68, df = 4 (p < 0.01)** |
| Abundance | 25 | -0.06 [-0.12; -0.00] |  |
| NDVI/EVI | 11 | -0.24 [-0.55; 0.07] |  |
| Canopy | 3 | -0.44 [-0.62; -0.27] |  |
| Visitation | 5 | 0.01 [0.00; 0.01] |  |
| Proximity | 1 | -0.02 [-0.08; 0.04] |  |
| **Covariates: demographic** | **45** | **-0.08 [-0.14; -0.03]** | **NA** |
| With demographic covariates | 45 | -0.08 [-0.14; -0.03] |  |
| Without demographic covariates | 0 |  |  |
| **Covariates: health** | **45** | **-0.08 [-0.14; -0.03]** | **1.35, df = 1 (p = 0.25)** |
| With health covariates | 37 | -0.06 [-0.11; 0.00] |  |
| Without health covariates | 8 | -0.13 [-0.28; 0.01] |  |
| **Covariates: economic** | **45** | **-0.08 [-0.14; -0.03]** | **NA** |
| With economic covariates | 45 | -0.08 [-0.14; -0.03] |  |
| Without economic covariates | 0 |  |  |
| **Covariates: climatic** | **45** | **-0.08 [-0.14; -0.03]** | **0.35, df = 1 (p = 0.56)** |
| With climatic covariates | 19 | -0.12 [-0.27; 0.03] |  |
| Without climatic covariates | 26 | -0.08 [-0.14; -0.02] |  |
| **Covariates: political** | **45** | **-0.08 [-0.14; -0.03]** | **8.27, df = 1 (p < 0.01)** |
| With political covariates | 14 | -0.01 [-0.04; 0.01] |  |
| Without political covariates | 31 | -0.15 [-0.24; -0.05] |  |

\*NDVI=Normalised Difference Vegetation Index; EVI= Enhanced Vegetation Index

### Greenness and COVID-19 mortalities

We found that an increase in greenness was strongly linked to lower mortality rate ratio (MRR= 0.9272; 95% CI: 0.8788 – 0.9783; t=-3.05; p=0.009) with a prediction interval of [0.7683 – 1.1189] (95% CI) (Figure 4). Furthermore, an estimated 0.0069 between-study heterogeneity variance (95% CI: 0.0032 – 0.0228) was observed with an I^2^ value of 92% (95% CI: 88.3% – 94.5%). We also found that year of publication (*X*^2^=19.10; p<0.01), sample size (*X*^2^=7.92; p<0.01), choice of predictors (*X*^2^=14.92; p<0.01), and use of health (*X*^2^=7.92; p<0.01) and political (*X*^2^=22.75; p<0.01) covariates strongly impact the degree of heterogeneity (see Table 2 & Figures S7 – S13).

**Figure 4.**
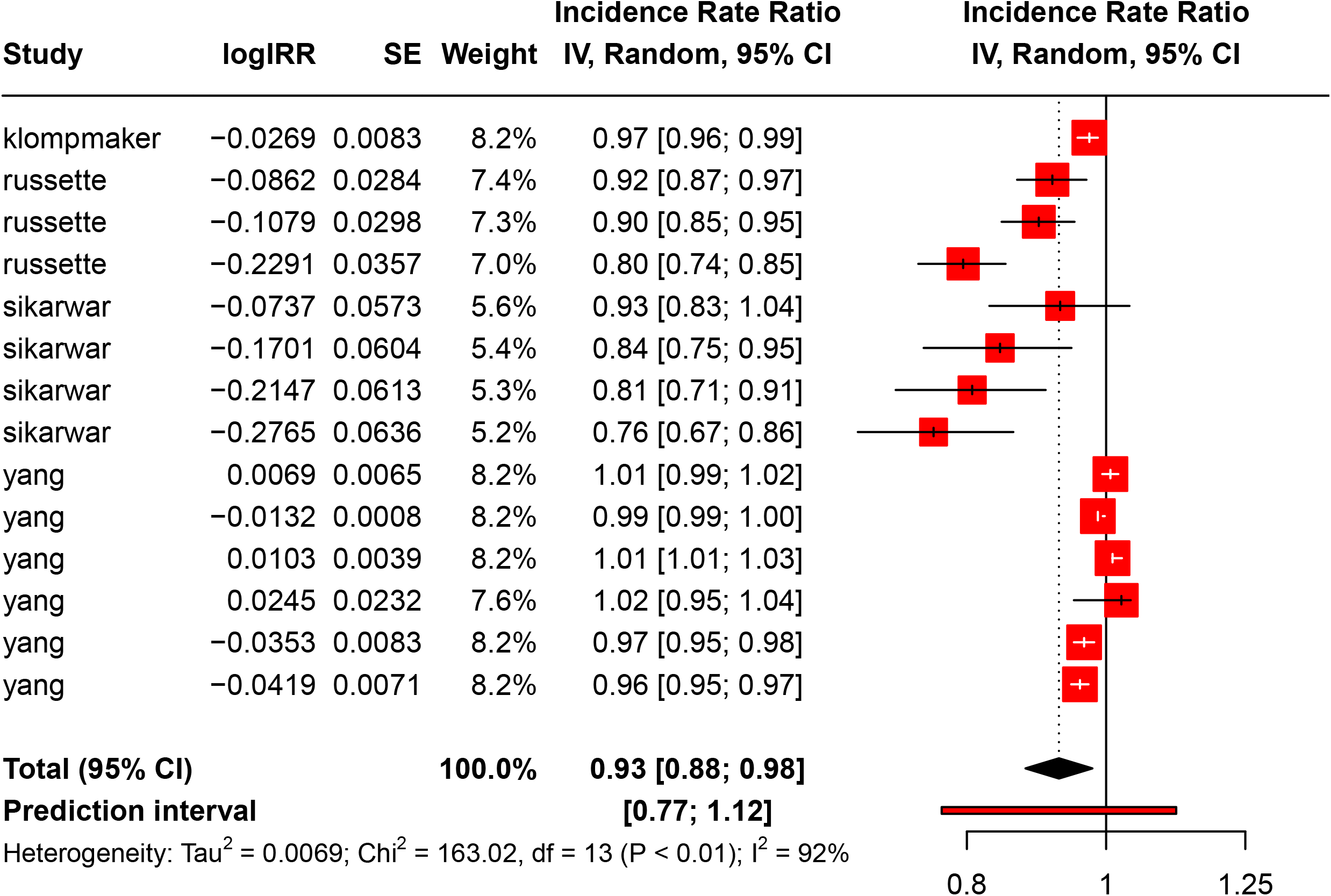
Forest plot of the relationship between greenness and COVID-19 mortalities.

**Table 2:** Stratified analyses of pooled mortality rate ratio of COVID-19 deaths.

| Stratified analysis | Number of results | Pooled mortality rate ratio [95% CI] | Subgroup difference $\chi^2$, df (p-value) |
| --- | --- | --- | --- |
| **Year of publication** | **14** | **0.93 [0.88; 0.98]** | **19.10, df = 2 (p< 0.01)** |
| 2021 | 4 | 0.90 [0.78; 1.03] |  |
| 2022 | 6 | 0.99 [0.96; 1.02] |  |
| 2023 | 4 | 0.83 [0.73; 0.96] |  |
| **Sample size** | **14** | **0.93 [0.88; 0.98]** | **7.92, df = 1 (p< 0.01)** |
| Small (n<2000) | 4 | 0.83 [0.73; 0.96] |  |
| Large (n≥2000) | 10 | 0.96 [0.91; 1.01] |  |
| **Predictor** | **14** | **0.93 [0.88; 0.98]** | **14.92, df = 2 (p< 0.01)** |
| Canopy | 3 | 0.87 [0.72; 1.05] |  |
| NDVI/EVI | 5 | 0.87 [0.76; 0.99] |  |
| Abundance | 6 | 0.99 [0.96; 1.02] |  |
| **Covariates: demographic** | **14** | **0.93 [0.88; 0.98]** | **NA** |
| With demographic covariates | 14 | 0.93 [0.88; 0.98] |  |
| Without demographic covariates | 0 |  |  |
| **Covariates: health** | **14** | **0.93 [0.88; 0.98]** | **7.92, df = 1 (p< 0.01)** |
| With health covariates | 10 | 0.96 [0.91; 1.01] |  |
| Without health covariates | 4 | 0.83 [0.73; 0.96] |  |
| **Covariates: economic** | **14** | **0.93 [0.88; 0.98]** | **2.60, df = 1 (p= 0.11)** |
| With economic covariates | 11 | 0.95 [0.89; 1.01] |  |
| Without economic covariates | 3 | 0.87 [0.72; 1.05] |  |
| **Covariates: climatic** | **14** | **0.93 [0.88; 0.98]** | **2.60, df = 1 (P = 0.11)** |
| With climatic covariates | 11 | 0.95 [0.89; 1.01] |  |
| Without climatic covariates | 3 | 0.87 [0.72; 1.05] |  |
| **Covariates: political** | **14** | **0.93 [0.88; 0.98]** | **22.75, df = 1 (p< 0.01)** |
| With political covariates | 7 | 0.85 [0.80; 0.92] |  |
| Without political covariates | 7 | 0.99 [0.97; 1.01] |  |

\*NDVI=Normalised Difference Vegetation Index; EVI= Enhanced Vegetation Index

### Publication bias

Existence of publication bias was investigated using the Funnel approach and Orwin fail-safe number. The presence of funnel plot symmetry (Figure 5A) indicated a lack of publication bias for studies that investigate the effect of greenness on COVID-19 infections (Fail-safe N: 45). Publication bias was, however, observed for studies that test the relationship between greenness and COVID-19 mortalities (Figure 5B; Fail-safe N: 14).

**Figure 5.**
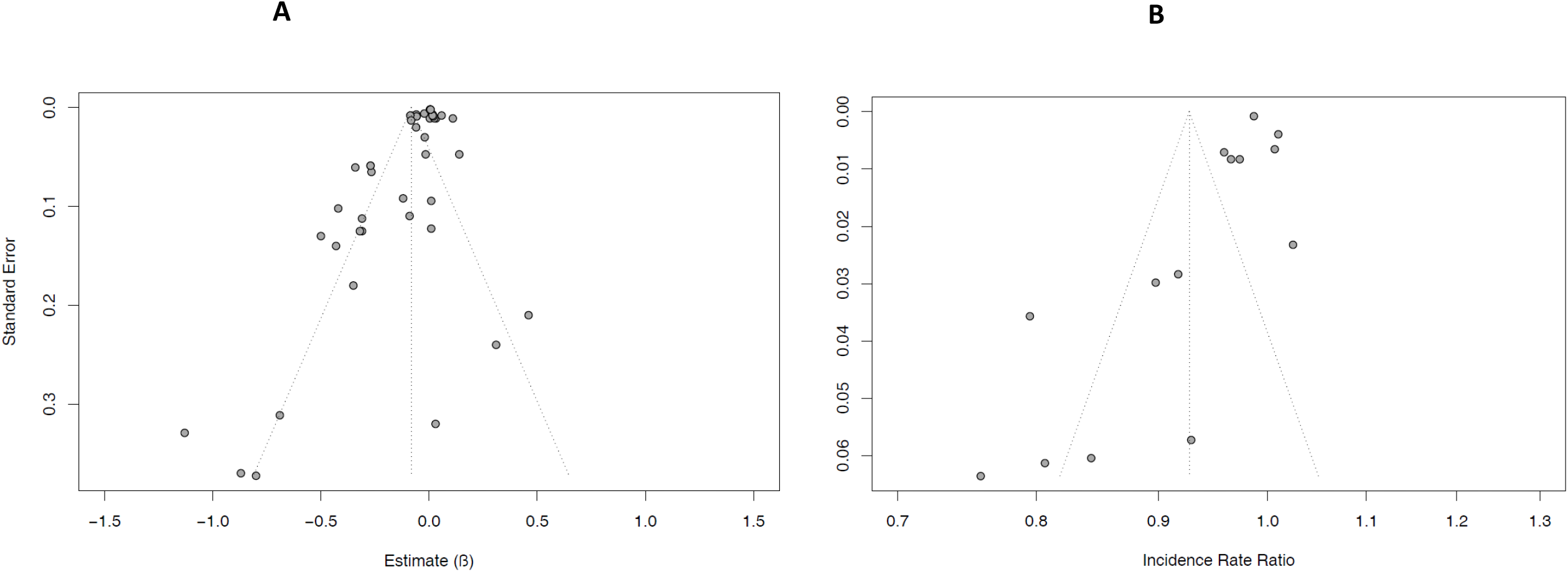
Funnel plot to test for publication bias in studies on A) COVID-19 infections, and B) on COVID-19 mortality.

## Discussion

Our meta-analysis provides evidence that an increase in abundance or exposure to greenness is associated with a significant reduction in COVID-19 infection rates and death cases^10,19,27^. However, we found high heterogeneity between the studies that were included in the meta-analysis. Subgroup analyses revealed that heterogeneity in studies on COVID-19 infections and mortality is strongly predicted by the studies’ years of publication, choices of predictors (metrics of greenness), and inclusion of political covariates. Additionally, sample size and consideration of health covariates strongly affect heterogeneity of studies on COVID-19 mortalities.

The sensitivity of effect size to year of publication can be attributed to availability of data to adequately model the impact of COVID-19. The spread of COVID-19 and increased global testing for COVID-19 infection accelerated overtime, thus allowing successive studies to have an increasingly larger data pool^28,29^. This may also impact sample sizes that are adopted in each study. As more regions produce more data on COVID-19 infections and mortality, their eligibility to be included in studies investigating the correlations between COVID-19 and greenness may enhance study designs. In our subgroup analysis, we found that studies that used smaller sample sizes (n<2000) are likely to report larger effect sizes compared to studies with larger sample sizes. Given the importance of selecting an appropriate sample size^30^, the need to define an appropriate sample size for investigating the health benefits of green infrastructure remains critical.

The diversity of greenness metrics, ranging from street trees to large forests, presents a unique challenge while measuring their impacts. Commonly, studies that cover large study areas use vegetation indices such as NDVI or EVI which are retrieved from satellite imagery^31-33^. Since health benefits of greenness are usually felt closer to the greenness^34,35^, several studies consider local greenness such as household gardens^36^, street trees^37,38^, and local parks^39,40^ in their analysis. However, this approach is only feasible when focusing on smaller areas. In some cases, subjective measures of greenness were used^41,42^. Our findings in the present study suggest that the choice of greenness metrics adopted in different studies affects the its effect size. The use of NDVI, EVI or vegetation canopy size produces large effects of greenness against COVID-19 infections and mortalities. In contrast, studies that use proximity or visitation patterns are likely to report marginal effects.

All studies in our meta-analysis have included demographic covariates, and 92% of studies included economic covariates. While modelling the effects of greenness, the inclusion of demographic variables such as population density and age structure, as well as economic indicators such as gross domestic product (GDP) and household income level as covariates have been largely adopted^7,9,27,43^. Furthermore, the use of health covariates featured in several studies^5,19,26^. However, consideration of political covariates in the modelling of greenness benefits to human wellbeing in the context of COVID-19 is only starting to emerge^5,6^. Political factors such as promulgation of mobility restrictions^15,44^ and face-masks mandates^45^ have shown to be significant predictors of COVID-19 impacts, although their inclusion in studies linking greenness to COVID-19 infection and severity remains limited. We found that the use of political covariates significantly affects the effect size. Inclusion of political covariables resulted in a greater effect size in studies of COVID-19 mortality and in a smaller effect size in studies of COVID-19 infections. This may suggest that existing policies are more effective in reducing COVID-19 fatalities than curbing the spread of infections.

Overall, meta-analysing studies from Africa, Asia, Europe, and USA, we found strong support for beneficial effects of greenness to human in the face of COVID-19 infection and severity, suggesting that positive correlations reported in some studies between greenness vs. infection and mortality rates^15,16^ might simply imply that the greenness metrics used in those studies (e.g., green space density or accessibility to greenspaces) do not fully capture important facets of greenness. This calls for a need to homogenize greenness metrics in studies to come. There is also a need for homogenization of COVID-19 severity metrics since we could not include hospitalization rate in the present study as a measure of COVID-19 severity because very limited studies have investigated hospitalization rate. Lastly, our results showed high degree of between-study heterogeneity which can be explained by year of publication, sample size, and choice of predictor variables and covariates. However, evidence from existing studies show that green infrastructure moderates the impact of COVID-19 by reducing prevalence of infections and associated mortalities.

Nevertheless, our findings have some far-reaching implications for the establishment and management of green infrastructure: greenspaces must be acknowledged as critical infrastructure that has substantial broader public health values, and as such, deserve enough fundings from governments worldwide, especially in the developing world.

## Methods

### Study selection

The Preferred Reporting Items for Systematic Reviews and Meta-Analyses (PRIMSA) guidelines^46^ were followed to search for literature that focus on green infrastructure and its impact on COVID-19. All search results were reviewed for relevance based on their title and abstract to be considered for meta-analysis (Figure 6). Furthermore, reference lists of all included articles were reviewed to identify studies that meet the inclusion criteria.

**Figure 6.**
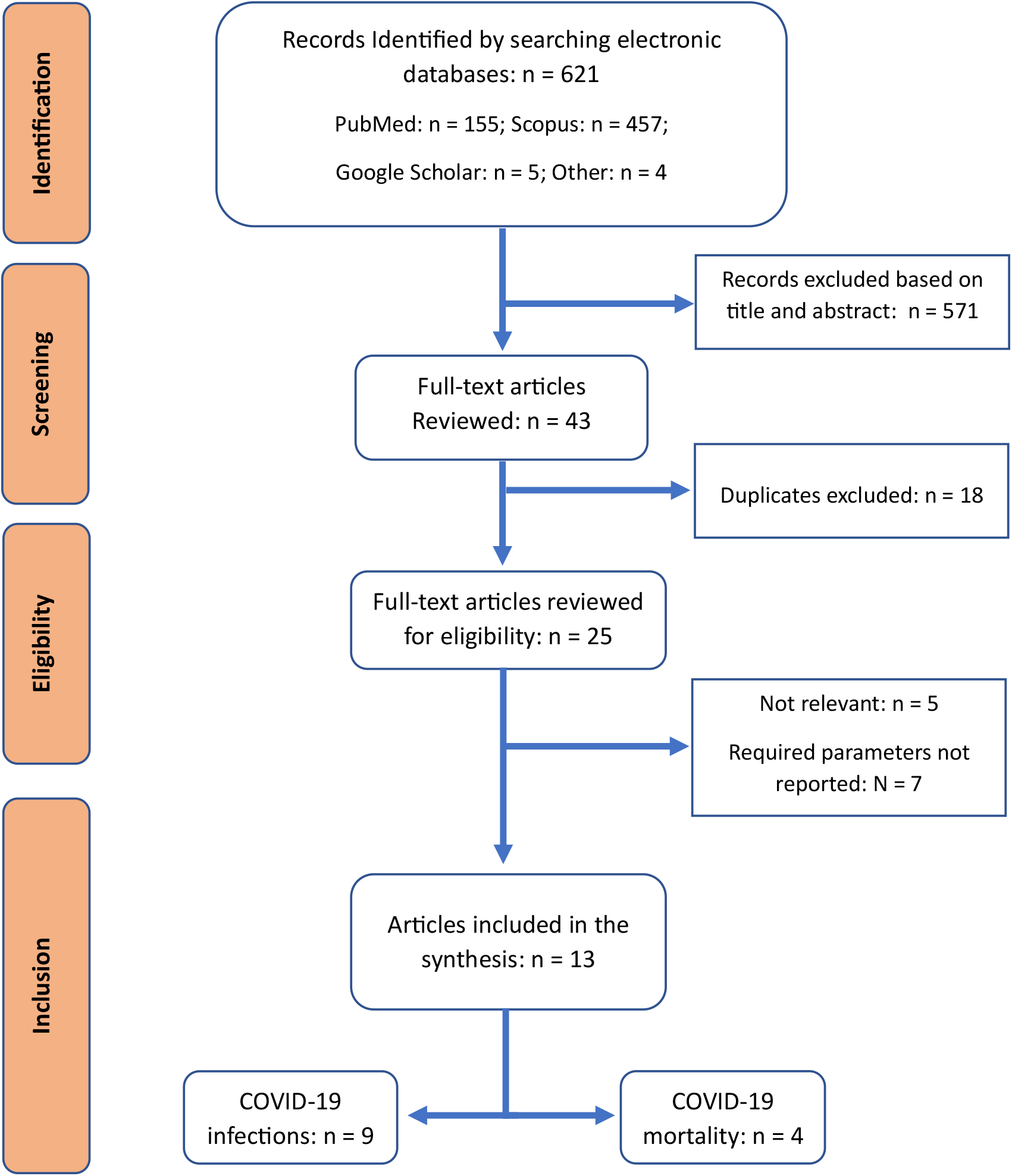
PRISMA low diagram for literature search and screening.

### Search strategy

Literature search was limited to PubMed, Scopus, and Google Scholar. The following search string was used to search for literature on the 17^th^ of April 2023: (“Greenspace“ or “green space“ or “greenery“ or “greenness“ or “vegetation“ or “trees“ or “forest” or “grass” or “grassland”) and (“COVID-19“ or “SARS-CoV-2“ or “coronavirus“ or “COVID”). We did not apply any restrictions on publication date in the search.

### Eligibility criteria

Inclusion criteria for this study were as follow: (a) original research that investigates effects of green infrastructure on COVID-19 infections and related mortalities; (b) full-text is available; (c) publication is in English; (d) required statistical parameters for meta-analysis are reported in the main article or supplementary files (i.e., regression estimates for predicting COVID-19 infections, and mortality rate ratios for predicting COVID-19 mortalities). Exclusion criteria were review or commentary articles, articles without required parameters, and articles not in English (see Figure 6).

### Data extraction and analysis

A predetermined template was used to collect study characteristics which are surname of first author, year of publication, country of study, measure of green infrastructure, temporal extent of study, sample size, measure of COVID-19, effect type, effect size, standard error or confidence interval, and list of covariates. All data analysed in this study are available as Supplemental Information (Appendix 1).

All analyses were conducted in R version 4.2.3^47^ (see R script in Supplemental Information). Two separate meta-analyses were conducted, focusing on impacts of greenness on COVID-19 infections (*meta-analysis 1*) and COVID-19 mortalities (*meta-analysis 2*). Regression estimates were used as pre-calculated effect type when analysing COVID-19 infections, and mortality rate ratios (MRR) were used as pre-calculated effect type when analysing COVID-19 mortalities. Subsequently, subgroup analyses were applied to the same data to test the effects of predictor variables, sample size, and selection of covariates. Random models were selected in each analysis using *metagen* function found in the “Metafor” R library^48^.

Outcomes are reported as pooled regression estimates for COVID-19 infections and as pooled MRR for COVID-19 deaths. Furthermore, in each case, a 95% confidence interval (CI), t-value, and p-values are reported with p<0.005 considered as an indicator of statistical significance. Between-studies heterogeneity was quantified using Higgins & Thompson’s I^2^ statistic^49^ with the I^2^ value of less than 25%, 50% and 75% indicating low, moderate, or high heterogeneity, respectively. Heterogeneity variance and prediction interval were also reported to measure the extent of between-study heterogeneity. Publication bias was tested using the Funnel approach^50^ (Sterne & Egger 2001) and the Orwin’s fail-safe number^51^.

## Data Availability

All data produced in the present work are contained in the manuscript.

https://figshare.com/s/207d839ca9d5ebe945fd

## Acknowledgements

KY acknowledges the South Africa’s National Research Foundation (grant # SRUG22051210107).

## Author Contributions

KY conceived the project, BP collected the data, BP analysed the data, KY and BP wrote the paper.

## Data Availability

All data generated or analysed in this study are included in this article (and its Supplementary Information files).

## Conflict of Interest Statement

The authors declare no conflict of interests.

## Code availability

Code to replicate all results in this paper is available as Supplemental Information.

## SUPPLEMENTAL INFORMATION

## TABLES

**Table S1**. Characteristics of all studies included in the present meta-analysis.

**Figures S1-S6**. Subgroup analyses of between-study heterogeneity can be attributed to year of publication (*X*^2^=8.24; p=0.02), choice of predictors (*X*^2^=129.68; p<0.01), and use of political covariates (*X*^2^=8.27; p<0.01).

**Figures S7-S13**. Between-study heterogeneity of variance showing that year of publication (*X*^2^=19.10; p<0.01), sample size (*X*^2^=7.92; p<0.01), choice of predictors (*X*^2^=14.92; p<0.01), and use of health (*X*^2^=7.92; p<0.01) and political (*X*^2^=22.75; p<0.01) covariates strongly impact the degree of heterogeneity.

**Appendices**

**Appendix 1**. Data collected and analysed in this study.

**Appendix 2**. R script used to reproduce the present study.

## Notes

### Competing Interest Statement

The authors have declared no competing interest.

### Funding Statement

This study was funded by the National Research Foundation South Africa

## References

1 UN DESA (United Nations Department of Economic and Social Affairs). Global Population Growth and Sustainable Development Goals. 2021. Available online: https://www.un.org/development/desa/pd/sites/www.un.org.development.desa.pd/files/undesa_pd_2022_global_population_growth.pdf (accessed on 12 December 2021).

2 Bongaarts, J. 2016. Development: Slow down population growth. Nature 530, 409–412.

3 WHO (World Health Organisation). 2020. Coronavirus disease 2019 (COVID-19), Situation Report– 65. Available at https://www.who.int/publications/m/item/situation-report---65

4 WHO (World Health Organisation). 2022. WHO coronavirus (COVID-19) dashboard. vailable at https://covid19.who.int/

5 Yang, Y., Lu, Y. & Jiang, B. (2022). Population-weighted exposure to green spaces tied to lower COVID-19 mortality rates: A nationwide dose-response study in the USA. Science of The Total Environment, 851: 158333. https://doi.org/10.1016/j.scitotenv.2022.158333.

6 Zhai, W., Yue, H. & Deng, Y. (2022). Examining the association between urban green space and viral transmission of COVID-19 during the early outbreak. Applied Geography, 147: 102768. https://doi.org/10.1016/j.apgeog.2022.102768.

7 Spotswood, E.N., Benjamin, M., Stoneburner, L., Wheeler, M. M., Beller, E. E., Balk, D., McPhearson, T., Kuo, M. & McDonald, R. I. (2021). Nature inequity and higher COVID-19 case rates in less-green neighbourhoods in the United States. Nature Sustainability, 4(12): 1092–1098. https://doi.org/10.1038/s41893-021-00781-9.

8 Nguyen, Q.C., Huang, Y., Kumar, A., Duan, H., Keralis, J.M., Dwivedi, P., Meng, H.W., Brunisholz, K.D., Jay, J., Javanmardi, M. & Tasdizen, T. (2020). Using 164 Million Google Street View Images to Derive Built Environment Predictors of COVID-19 Cases. International Journal of Environmental Research and Public Health, 17(17): 6359. Doi: 10.3390/ijerph17176359. PMID: 32882867; PMCID: PMC7504319.

9 Peng, W., Dong, Y., Tian, M., Yuan, J., Kan, H., Jia, X. & Wang, W. (2022). City-level greenness exposure is associated with covid-19 incidence in China. Environmental Research, 209: 112871. https://doi.org/10.1016/j.envres.2022.112871.

10 Sikarwar, A., Rani, R., Duthé, G. & Golaz, V. (2023). Association of Greenness with covid-19 deaths in India: An Ecological Study at district level. Environmental Research, 217: 114906. https://doi.org/10.1016/j.envres.2022.114906.

11 Li, Q. 2010. Effects of forest bathing trips on human immune function. Environmental Health and Preventive Medicine, 15, 9–17.

12 Vivier, E., Tomasello, E., Baratin, M., Walzer, T. & Ugolini, S. 2008. Functions of natural killer cells. Nature Immunology, 9, 503–510.

13 Chen, Q.X., Huang, C.L., Yuan, Y. & Tan, H.P. 2020. Influence of COVID-19 event on air quality and their association in mainland China. Aerosol and Air Quality Research, 20: 1541–1551.

14 Lin, J., Kroll, C. N., Nowak, D. J., & Greenfield, E. J. (2019). A review of urban forest modeling: Implications for management and future research. Urban Forestry & Urban Greening, 43, Article 126366

15 Huang, X., Shao, X., Xing, L., Hu, Y., Sin, D.D. & Zhang, X. (2021). The impact of lockdown timing on COVID-19 transmission across US counties. EClinicalMedicine, 38: 101035. https://doi.org/10.1016/j.eclinm.2021.101035.

16 Pan J., Bardhan R., Jin Y. ^*^, Ying Jin 2021. Spatial distributive effects of public green space and COVID-19 infectionin London

17 Zhang, L. & Tan, P.Y. (2019). Associations between Urban Green Spaces and Health are Dependent on the Analytical Scale and How Urban Green Spaces are Measured. International Journal of Environmental Research and Public Health, 16(4): 578. https://doi.org/10.3390/ijerph16040578

18 Labib S.M., Lindley S. & Huck J.J. (2020). Scale effects in remotely sensed greenspace metrics and how to mitigate them for environmental health exposure assessment. Computers, Environment and Urban Systems, 82: 101501. Doi: 10.1016/j.compenvurbsys.2020.101501.

19 Jiang, B., Yang, Y., Chen, L., Liu, X., Wu, X., Chen, B., Webster, C., Sullivan, W. C., Larsen, L., Wang, J., & Lu, Y. (2022). Green spaces, especially nearby forest, may reduce the SARS-COV-2 infection rate: A nationwide study in the United States. Landscape and Urban Planning, 228: 104583. https://doi.org/10.1016/j.landurbplan.2022.104583.

20 Chen, K., Klompmaker, J. O., Roscoe, C. J., Nguyen, L. H., Drew, D. A., James, P., Laden, F., Fecht, D., Wang, W., Gulliver, J., Wolf, J., Steves, C. J., Spector, T. D., Chan, A. T. & Hart, J. E. (2023). Associations between greenness and predicted COVID-19-like illness incidence in the United States and the United Kingdom. Environmental Epidemiology (Philadelphia, Pa.), 7(1): e244. https://doi.org/10.1097/EE9.0000000000000244.

21 Bajaj, V., Gadi, N., Spihlman, A. P., Wu, S. C., Choi, C. H. & Moulton, V. R. (2021). Aging, Immunity, and COVID-19: How Age Influences the Host Immune Response to Coronavirus Infections? Frontiers in Physiology, 11. https://doi.org/10.3389/fphys.2020.571416.

22 Mathur, R., Rentsch, C.T., Morton, C.E., Hulme, W.J., Schultze, A., Mackenna, B., Eggo, R.M., Bhaskaran, K., Wong, A.Y.S., Williamson, E.J., Forbes, H., Wing, K., Mcdonald, H.I., Bates, C., Bacon, S., Walker, A.J., Evans, D., Inglesby, P., Mehrkar, A., Curtis, H.J., Devito, N.J., Croker, R., Drysdale, H., Cockburn, J., Parry, J., Hester, F., Harper, S., Douglas, I.J., Tomlinson, L., Evans, S.J.W., Grieve, R., Harrison, D., Rowan, K., Khunti, K., Chaturvedi, N., Smeeth, L. & Goldacre, B. (2021). Ethnic differences in SARS-CoV-2 infection and COVID-19-related hospitalisation, intensive care unit admission, and death in 17 million adults in England: an observational cohort study using the OpenSAFELY platform. The Lancet, 397(10286): 1711 – 1724.

23 Hussey, H., Zinyakatira, N., Morden, E., Ismail, M., Paleker, M., Bam, L., London, L., Boulle, A. & Davies, A. (2021). Higher COVID-19 mortality in low-income communities in the City of Cape Town – a descriptive ecological study. Gates Open Research, 5. https://doi.org/10.12688/gatesopenres.13288.1.

24 Lu, Y., Chen, L., Liu, X., Yang, Y., Sullivan, W. C., Xu, W., Webster, C. & Jiang, B. (2021). Green spaces mitigate racial disparity of health: A higher ratio of green spaces indicates a lower racial disparity in SARS-COV-2 infection rates in the USA. Environment International, 152: 106465. https://doi.org/10.1016/j.envint.2021.106465.

25 Li, H., Zhang, G. & Cao, Y. (2022). Forest Area, CO2 Emission, and COVID-19 Case-Fatality Rate: A Worldwide Ecological Study Using Spatial Regression Analysis. Forests, 13(5), 736. https://doi.org/10.3390/f13050736.

26 Lin, J., Huang, B., Kwan, M.P., Chen, M. & Wang, Q. (2023). Covid-19 infection rate but not severity is associated with availability of greenness in the United States. Landscape and Urban Planning, 233: 104704. https://doi.org/10.1016/j.landurbplan.2023.104704.

27 Klompmaker, J.O., Hart, J.E., Holland, I., Sabath, M.B., Wu, X., Laden, F., Dominici, F. & James, P. (2020). County-level exposures to Greenness and associations with covid-19 incidence and mortality in the United States. Science of the Total Environment, 199: 111331. https://doi.org/10.1101/2020.08.26.20181644.

28 Singh, S., Chowdhury, C., Panja, A. K. & Neogy, S. (2021). Time Series Analysis of COVID-19 Data to Study the Effect of Lockdown and Unlock in India. Journal of The Institution of Engineers (India): Series B, 102(6): 1275–1281. https://doi.org/10.1007/s40031-021-00585-7

29 OWD (Our World in Data). 2023. Total COVID-19 tests. https://ourworldindata.org/grapher/full-list-total-tests-for-covid-19. (Accessed 02.05.2023).

30 Springate, S. D. (2012). The effect of sample size and bias on the reliability of estimates of error: A comparative study of Dahlberg’s formula. European Journal of Orthodontics, 34(2): 158–163. https://doi.org/10.1093/ejo/cjr010

31 Fong, K., Hart, J. E. & James, P. (2018). A Review of Epidemiologic Studies on Greenness and Health: Updated Literature Through 2017. Current environmental health reports, 5(1): 77. https://doi.org/10.1007/s40572-018-0179-y.

32 Brochu, P., Jimenez, M. P., James, P., Kinney, P. L. & Lane, K. (2022). Benefits of Increasing Greenness on All-Cause Mortality in the Largest Metropolitan Areas of the United States Within the Past Two Decades. Frontiers in Public Health, 10. https://doi.org/10.3389/fpubh.2022.841936.

33 Grigsby-Toussaint, D.S. & Shin, J.C. (2022). Covid-19, Green Space Exposure, and Mask Mandates. Science of The Total Environment, 836: 155302. https://doi.org/10.1016/j.scitotenv.2022.155302.

34 Ngom, R., Gosselin, P., Blais, C. & Rochette, L. (2016). Type and Proximity of Green Spaces Are Important for Preventing Cardiovascular Morbidity and Diabetes—A Cross-Sectional Study for Quebec, Canada. International Journal of Environmental Research and Public Health, 13(4). https://doi.org/10.3390/ijerph13040423.

35 Dennis, M., Cook, P.A., James, P., Wheater, P. & Lindley, S.J. (2020). Relationships between health outcomes in older populations and urban green infrastructure size, quality and proximity. BMC Public Health 20, 626. https://doi.org/10.1186/s12889-020-08762-x.

36 Chalmin-Pui, L. S., Griffiths, A., Roe, J., Heaton, T. & Cameron, R. (2021). Why garden? – Attitudes and the perceived health benefits of home gardening. Cities, 112: 103118. https://doi.org/10.1016/j.cities.2021.103118.

37 Marselle, M. R., Bowler, D. E., Watzema, J., Eichenberg, D., Kirsten, T., & Bonn, A. (2020). Urban street tree biodiversity and antidepressant prescriptions. Scientific Reports, 10(1), 1–11. https://doi.org/10.1038/s41598-020-79924-5.

38 Wolf, K. L., Lam, S. T., McKeen, J. K., Richardson, R. A. & Bardekjian, A. C. (2020). Urban Trees and Human Health: A Scoping Review. International Journal of Environmental Research and Public Health, 17(12). https://doi.org/10.3390/ijerph17124371

39 Orstad, S. L., Szuhany, K., Tamura, K., Thorpe, L. E. & Jay, M. (2020). Park Proximity and Use for Physical Activity among Urban Residents: Associations with Mental Health. International Journal of Environmental Research and Public Health, 17(13): 4885. https://doi.org/10.3390/ijerph17134885.

40 Weber, K.A., Yang, W., Carmichael, S. L., Collins, R.T., Luben, T.J., Desrosiers, T.A., Insuf, T.I., Le, M.T., Evans, S.P., Romitti, P.A., Yazdy, M.M., Nembhard, W.N. & Shaw, G.M. (2023). The National Birth Defects Prevention Study. (2023). Assessing associations between residential proximity to greenspace and birth defects in the national birth defects prevention study. Environmental Research, 216: 114760 doi:10.1016/j.envres.2022.114760.

41 Yessoufou, K., Sithole, M. & Elansary, H. O. (2020). Effects of urban green spaces on human perceived health improvements: Provision of green spaces is not enough but how people use them matters. PLOS ONE, 15(9): e0239314. https://doi.org/10.1371/journal.pone.0239314.

42 Lehberger, M., Kleih, A. & Sparke, K. (2021). Self-reported well-being and the importance of green spaces – A comparison of garden owners and non-garden owners in times of COVID-19. Landscape and Urban Planning, 212, 104108. https://doi.org/10.1016/j.landurbplan.2021.104108.

43 Russette, H., Graham, J., Holden, Z., Semmens, E.O., Williams, E. & Landguth, E.L. (2021). Greenspace exposure and covid-19 mortality in the United States: January–July 2020. Environmental Research, 198: 111195. https://doi.org/10.1016/j.envres.2021.111195.

44 Haug, N., Geyrhofer, L., Londei, A., Dervic, E., Loreto, V., Pinior, B., Thurner, S. & Klimek, P. (2020). Ranking the effectiveness of worldwide COVID-19 government interventions. Nature Human Behaviour, 4(12): 1303–1312. https://doi.org/10.1038/s41562-020-01009-0.

45 Aravindakshan, A., Boehnke, J., Gholami, E. & Nayak, A. (2022). The impact of mask-wearing in mitigating the spread of COVID-19 during the early phases of the pandemic. PLOS Global Public Health 2(9): e0000954. https://doi.org/10.1371/journal.pgph.0000954.

46 Page, M.J., McKenzie, J.E., Bossuyt, P.M., Boutron, I., Hoffmann, T.C., Mulrow, C.D., Shamseer, L., Tetzlaff, J.M., Akl, E.A., Brennan, S.E., Chou, R., Glanville, J., Grimshaw, J.M., Hróbjartsson, A., Lalu, M.M., Li, T., Loder, E.W., Mayo-Wilson, E., McDonald, S., McGuinness, L.A., Stewart, L.A., Thomas, J., Tricco, A.C., Welch, V.A., Whiting, P. & Moher, D. (2021). The PRISMA 2020 statement: An updated guideline for reporting systematic reviews. PLoS Med 18(3): e1003583. https://doi.org/10.1371/journal.pmed.1003583.

47 R Core Team. (2021). R: A language and environment for statistical computing. R Foundation for Statistical Computing, Vienna, Austria. https://www.R-project.org/.

48 Viechtbauer, W. (2010). Conducting meta-analyses in R with the metafor package. Journal of Statistical Software, 36(3): 1–48. https://doi.org/10.18637/jss.v036.i03.

49 Higgins, J.P. & Thompson, S.G. (2002). Quantifying heterogeneity in a meta-analysis. Statistics in Medicine, 21(11): 1539 – 1558. https://doi.org/10.1002/sim.1186.

50 Sterne, J.A.C. & Egger, M. (2001). Funnel plots for detecting bias in meta-analysis: guidelines on choice of axis. Journal of Clinical Epidemiology, 54:1046–1055

51 Orwin RG. A fail-safe N for effect size in meta-analysis. Journal of Educational Statistics. 1983;8(2):157–159.

