## Supplemental Table 1 for "A global meta-analysis of effects of green infrastructure on COVID-19 infection and mortality rates"

**Table 1**. Study characteristics.

| **Study**  **(1^st^ author, Year)** | **Country** | **Measure of green infrastructure** | **Temporal extent** | **Sample size** | **Measure of COVID-19 impact** | **Covariates** |
| --- | --- | --- | --- | --- | --- | --- |
| Grigsby-Toussaint, 2022 | United States of America | - Normalised Difference Vegetation Index - Tree Canopy | - 1 October 2020 | 3108 | Positive cases per 1000 people | - Rural population; Total population; Socioeconomic status; Household composition & disability; Minority status & language; Housing & transportation; Particulate matter (PM2.5); Precipitation; Temperature; Wind speed. |
| Jiang, 2022 | United States of America | - open space inside park, - open space outside park, - forest inside park, - forest outside park, - shrub and scrub, - herbaceous, - hay and pasture | 22 January - 31 December 2020 | 3108 | Infection rate | - Socioeconomic & demographic factors; Healthcare & testing factors; Pre-existing chronic disease factors; Politics & policy factors; Behavioural factors; Environmental Factors. |
| Johnson, 2021 | England | - median frequency of parks within a 1km^2^ radius around households - available green space per person (m^2^) within the local authority | 1 March 2020 – 30 November 2020 | 299 | Infection rate | - Lag case rate; Population density; Population clustering; Mobility; Case average; Baseline health; Percentage over 70; Percentage unemployed. |
| Klompmaker, 2021 | United States of America | - Mean values of Normalised Difference Vegetation Index | 1 April 2020 – 31 May 2020 | 3089 | COVID-19 death rate (per 100 000) | - Population density; % poverty; % owner occupied housing; % less than high school education; % black; % Hispanic; % 65+ years of age; % 45 – 64 years of age; %14 – 44 years of age; Median home value; Median household income; % obese; % current smokers; Days since stay-at-home order; Days since non-essential businesses closure; Days since nursing homes visitor ban; Days since first case; Rate of hospital beds; Rate of tests; Average summer temperature; Average winter temperature; Average summer relative humidity; Average winter relative humidity; PM2.5; Urban counties; Counties with issuance of stay-at-home order; Counties with 10< cases. |
| Lin, 2023 | United States of America | population-density-weighted NDVI | - April 2021 | 3040 | Count of COVID-19 infections | - Particulate matter (PM2.5); Temperature; % unemployment; % poverty; GINI; % not proficient in English; % bachelor; % females; % white; % 60 and older; % below 18; % rent; Average household size; % severe housing problems; % children in single-parent households; % limited access to healthy food; Social associations; % adult obesity; % adult diabetes; % physical inactivity; %excessive drinking. |
| Lu, 2021 | United States of America | - Developed open space - Forest - Shrub & scrub - Grassland & herbaceous - Pasture & hay - Cultivated crops - Woody wetlands - Emergent herbaceous wetlands | 10 July 2020 | 135 | Number of COVID-19 cases per 100 000 people | - Population density; Female population ratio; Different in black-white population; Different in black-white adult population; Household size; Households with broadband; Median household income; Healthcare receipts; Number of firms; coronary heart disease; death rate; Heart failure death rate; Diagnosed diabetes rate. |
| Peng, 2022 | China | - Mean value of Normalised Difference Vegetation Index | 1 January 2020 – 29 February 2020 | 266 | Number of COVID-19 cases per 100 000 people | - Population density; Older people %; Gender ratio; Education years; Urbanisation rate; GDP per capita; Hospital beds; Number of doctors; Government Response Index; Intra-city movement intensity; PM2.5; NO2; CO; Temperature; Relative humidity |
| Phogole, 2023 | South Africa | - Mean value of Enhanced Vegetation Index - Forest - Grassland | 4 June 2022 | 4429  226 | Count number of COVID-19 cases per total population in a unit area | - Aged 65 years or older; Surface area; Revenue per capita |
| Russette, 2021 | United states of South Africa | - Leaf area index | 21 January 2020 – 29 July 2020 | 3049 | Count number of COVID-19 deaths | - Total population; Over 60 %; No high school diploma or equivalent; Medical aid; Overcrowding; Black %; Native American %; Physical inactivity |
| Sikarwar, 2023 | India | - Normalised Difference Vegetation Index | - 1 May 2020 | 640 | Count number of COVID-19 deaths | - PM2.5; Temperature & rainfall; Total population; Population density; Proportion of older adults & their sex ratio; Rural population; Household crowding; Material deprivation |
| Spotswood, 2021 | United States of America | - Normalised Difference Vegetative Index - Park proximity | 1 – 30 September 2020 | 2652 | Number of COVID-19 cases per 100 000 people | - Proportion of non-white people; Income; Age; Population density; Days since first case |
| Yang, 2022 | United States of America | - Forest inside park - Forest outside park - Grassland/Herbaceous - Pasture/hay - Open space inside park - Open space outside park | 22 January 2020 – 31 December 2020 | 3025 | Number of COVID-19 deaths per 100 000 | - Population density; Black non-Hispanic; Population aged 65+; Gini index of income inequality; Median home value; Unemployment rate; Without high school diploma; Population without insurance; SARS-CoV-2 testing rate; Diabetes; Obesity; Stroke; Hypertension; Heart stroke; Smoker; Essential worker; Places of interest visits; Commute by walking; Physical inactivity; Mobility; Mobility index; Stay-at-hope orders; Public mask mandate; Bars closed and reopened; Restaurant closed and reopened; Crowded housing; Proximity to highway; Airport density; Railway density; Road density; PM2.5; PM10; NO2; Maximum temperature ; Humidity; Wind speed |
| Zhai, 2022 | United States of America | - Urban green spaces visitation | 27 February 2020 – 27 May 2020 | 3108 | Effective COVID-19 reproduction number | - Age; Proportion of blacks; Poverty rate; Population density; Essential occupation rate; Trump share; Healthcare workers |
