## Supplemental Figure for "A global meta-analysis of effects of green infrastructure on COVID-19 infection and mortality rates": Figure S1.pdf

| Study or Subgroup | Estimate (ß) | SE | Weight | Estimate (ß) IV, Random, 95% CI |
|---|---|---|---|---|
| year = 2021 | | | | |
| johnson | 0.1100 | 0.0110 | 3.0% | 0.11 [−0.01; 0.03] |
| johnson | 0.0035 | 0.0110 | 3.0% | 0.00 [−0.02; 0.02] |
| johnson | −0.0570 | 0.0084 | 3.0% | −0.06 [−0.07; −0.04] |
| johnson | 0.0320 | 0.0110 | 3.0% | 0.03 [ 0.01; 0.05] |
| johnson | 0.0240 | 0.0108 | 3.0% | 0.02 [ 0.00; 0.04] |
| lu | −0.3100 | 0.1250 | 1.6% | −0.31 [−0.56; −0.07] |
| lu | −0.3100 | 0.1122 | 1.8% | −0.31 [−0.53; −0.09] |
| lu | −0.3200 | 0.1250 | 1.6% | −0.32 [−0.56; −0.07] |
| lu | −0.4200 | 0.1020 | 1.9% | −0.42 [−0.62; −0.22] |
| lu | −0.1200 | 0.0918 | 2.1% | −0.12 [−0.30; 0.06] |
| lu | 0.0100 | 0.1225 | 1.6% | 0.01 [−0.23; 0.25] |
| lu | 0.0100 | 0.0944 | 2.0% | 0.01 [−0.18; 0.19] |
| lu | −0.0900 | 0.1097 | 1.8% | −0.09 [−0.30; 0.13] |
| spotswood | −0.0600 | 0.0200 | 2.9% | −0.06 [−0.10; −0.02] |
| spotswood | −0.0200 | 0.0300 | 2.9% | −0.02 [−0.08; 0.04] |
| Total (95% CI) | | | **35.1%** | **−0.07 [−0.15; 0.01]** |
| Heterogeneity: $Tau^2 = 0.0143$; $Chi^2 = 207.16$, df = 14 ($P < 0.01$); $I^2 = 93\%$ | | | | |
| year = 2022 | | | | |
| grigsby_toussaint | 0.3100 | 0.2400 | 0.7% | 0.31 [−0.16; 0.78] |
| grigsby_toussaint | 0.4600 | 0.2100 | 0.9% | 0.46 [ 0.05; 0.87] |
| grigsby_toussaint | 0.0300 | 0.3200 | 0.5% | 0.03 [−0.60; 0.66] |
| grigsby_toussaint | −0.4300 | 0.1400 | 1.4% | −0.43 [−0.70; −0.16] |
| grigsby_toussaint | −0.5000 | 0.1300 | 1.6% | −0.50 [−0.75; −0.25] |
| grigsby_toussaint | −0.3500 | 0.1800 | 1.1% | −0.35 [−0.70; 0.00] |
| jiang | −0.0590 | 0.0070 | 3.0% | −0.06 [−0.07; −0.05] |
| jiang | 0.0190 | 0.0090 | 3.0% | 0.02 [ 0.00; 0.04] |
| jiang | 0.0170 | 0.0070 | 3.0% | 0.02 [ 0.00; 0.03] |
| jiang | −0.0220 | 0.0060 | 3.0% | −0.02 [−0.03; −0.01] |
| jiang | 0.0160 | 0.0080 | 3.0% | 0.02 [ 0.00; 0.03] |
| jiang | 0.0580 | 0.0080 | 3.0% | 0.06 [ 0.04; 0.07] |
| jiang | −0.0580 | 0.0090 | 3.0% | −0.06 [−0.08; −0.04] |
| jiang | −0.0870 | 0.0080 | 3.0% | −0.09 [−0.10; −0.07] |
| peng | −0.0830 | 0.0130 | 3.0% | −0.08 [−0.11; −0.06] |
| zhai | 0.0040 | 0.0020 | 3.0% | 0.00 [ 0.00; 0.01] |
| zhai | 0.0050 | 0.0020 | 3.0% | 0.00 [ 0.00; 0.01] |
| zhai | 0.0060 | 0.0020 | 3.0% | 0.01 [ 0.00; 0.01] |
| zhai | 0.0060 | 0.0020 | 3.0% | 0.01 [ 0.00; 0.01] |
| zhai | 0.0070 | 0.0020 | 3.0% | 0.01 [ 0.00; 0.01] |
| Total (95% CI) | | | **48.0%** | **−0.02 [−0.06; 0.02]** |
| Heterogeneity: $Tau^2 = 0.0022$; $Chi^2 = 413.64$, df = 19 ($P < 0.01$); $I^2 = 95\%$ | | | | |
| year = 2023 | | | | |
| lin | −1.1300 | 0.3291 | 0.4% | −1.13 [−1.78; −0.49] |
| lin | −0.6900 | 0.3112 | 0.5% | −0.69 [−1.30; −0.08] |
| lin | −0.8700 | 0.3699 | 0.4% | −0.87 [−1.59; −0.14] |
| lin | −0.8000 | 0.3725 | 0.4% | −0.80 [−1.53; −0.07] |
| phogole | −0.2707 | 0.0588 | 2.5% | −0.27 [−0.39; −0.16] |
| phogole | −0.2665 | 0.0651 | 2.4% | −0.27 [−0.39; −0.14] |
| phogole | −0.0158 | 0.0473 | 2.7% | −0.02 [−0.11; 0.08] |
| phogole | −0.2707 | 0.0588 | 2.5% | −0.27 [−0.39; −0.16] |
| phogole | 0.1397 | 0.0473 | 2.7% | 0.14 [ 0.05; 0.23] |
| phogole | −0.3410 | 0.0605 | 2.5% | −0.34 [−0.46; −0.22] |
| Total (95% CI) | | | **16.9%** | **−0.32 [−0.57; −0.07]** |
| Heterogeneity: $Tau^2 = 0.0780$; $Chi^2 = 85.74$, df = 9 ($P < 0.01$); $I^2 = 90\%$ | | | | |
| **Total (95% CI)** | | | **100.0%** | **−0.08 [−0.14; −0.03]** |
| **Prediction interval** | | | | **[−0.36; 0.20]** |
| Heterogeneity: $Tau^2 = 0.0184$; $Chi^2 = 749.60$, df = 44 ($P < 0.01$); $I^2 = 94\%$ | | | | |
| Test for subgroup differences: $Chi^2 = 8.24$, df = 2 ($P = 0.02$) | | | | |

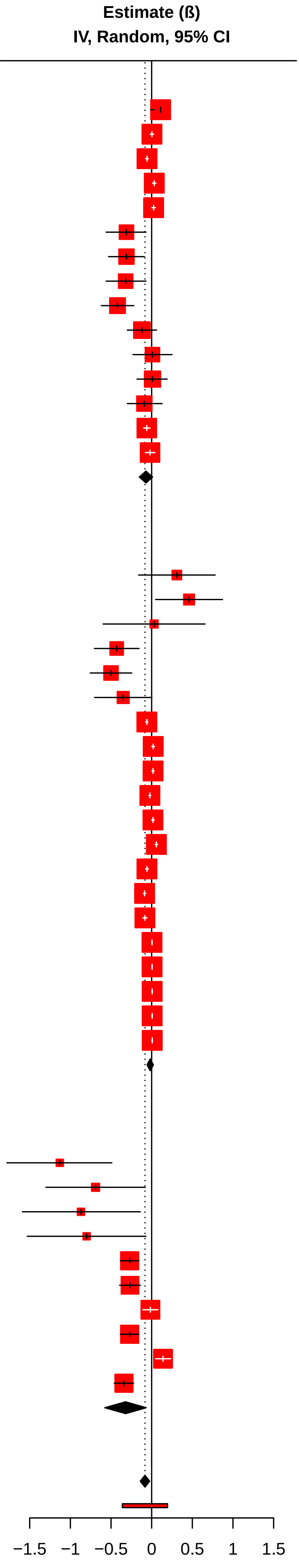
