## Supplemental Figure for "A global meta-analysis of effects of green infrastructure on COVID-19 infection and mortality rates": Figure S4.pdf

Heterogeneity: $Tau^2 = 0.0184$; $Chi^2 = 749.60$, df = 44 ($P < 0.01$); $I^2 = 94\%$

Test for subgroup differences: $Chi^2 = 1.35$, df = 1 ($P = 0.25$)

Estimate (ß) IV, Random, 95% CI

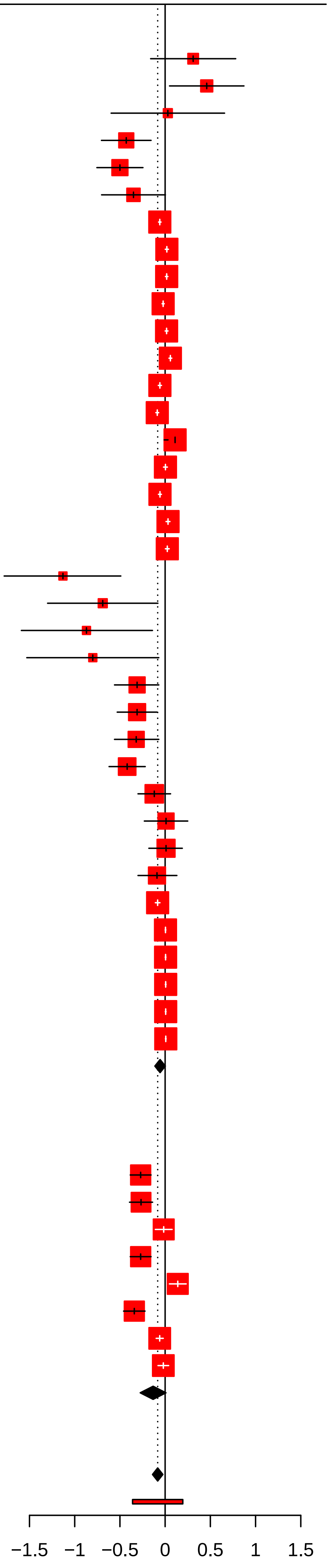
