## Supplemental Figure for "A global meta-analysis of effects of green infrastructure on COVID-19 infection and mortality rates": Figure S6.pdf

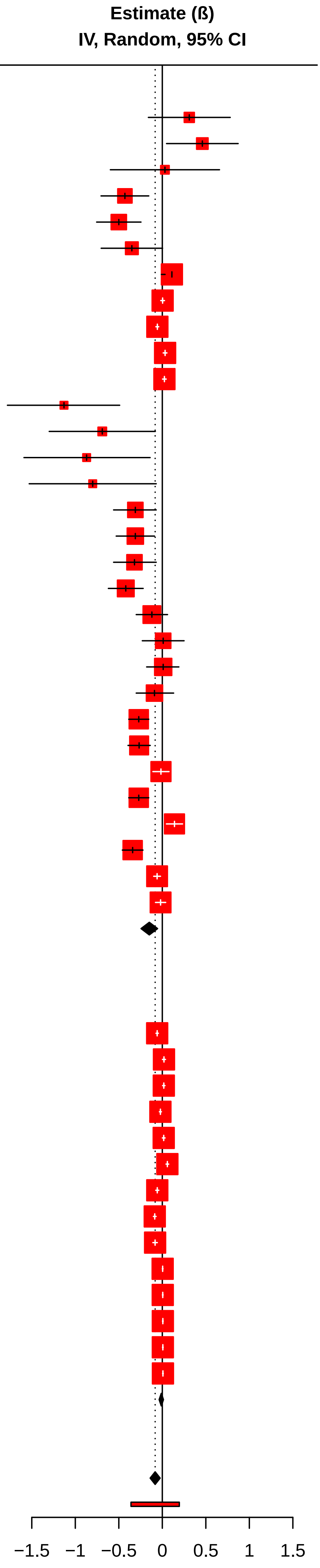
