## Supplemental Figure for "A global meta-analysis of effects of green infrastructure on COVID-19 infection and mortality rates": Figure S7.pdf

| Study or Subgroup | logIRR | SE | Weight | Incidence Rate Ratio IV, Random, 95% CI |
|---|---|---|---|---|
| year = 2021 | | | | |
| klompmaker | −0.0269 | 0.0083 | 8.2% | 0.97 [0.96; 0.99] |
| russette | −0.0862 | 0.0284 | 7.4% | 0.92 [0.87; 0.97] |
| russette | −0.1079 | 0.0298 | 7.3% | 0.90 [0.85; 0.95] |
| russette | −0.2291 | 0.0357 | 7.0% | 0.80 [0.74; 0.85] |
| **Total (95% CI)** | | | **29.8%** | **0.90 [0.78; 1.03]** |
| Heterogeneity: $Tau^2 = 0.0063$; $Chi^2 = 37.96$, df = 3 ($P < 0.01$); $I^2 = 92\%$ | | | | |
| year = 2023 | | | | |
| sikarwar | −0.0737 | 0.0573 | 5.6% | 0.93 [0.83; 1.04] |
| sikarwar | −0.1701 | 0.0604 | 5.4% | 0.84 [0.75; 0.95] |
| sikarwar | −0.2147 | 0.0613 | 5.3% | 0.81 [0.71; 0.91] |
| sikarwar | −0.2765 | 0.0636 | 5.2% | 0.76 [0.67; 0.86] |
| **Total (95% CI)** | | | **21.5%** | **0.83 [0.73; 0.96]** |
| Heterogeneity: $Tau^2 = 0.0038$; $Chi^2 = 6.1$, df = 3 ($P = 0.11$); $I^2 = 51\%$ | | | | |
| year = 2022 | | | | |
| yang | 0.0069 | 0.0065 | 8.2% | 1.01 [0.99; 1.02] |
| yang | −0.0132 | 0.0008 | 8.2% | 0.99 [0.99; 1.00] |
| yang | 0.0103 | 0.0039 | 8.2% | 1.01 [1.01; 1.03] |
| yang | 0.0245 | 0.0232 | 7.6% | 1.02 [0.95; 1.04] |
| yang | −0.0353 | 0.0083 | 8.2% | 0.97 [0.95; 0.98] |
| yang | −0.0419 | 0.0071 | 8.2% | 0.96 [0.95; 0.97] |
| **Total (95% CI)** | | | **48.7%** | **0.99 [0.96; 1.02]** |
| Heterogeneity: $Tau^2 = 0.0005$; $Chi^2 = 70.53$, df = 5 ($P < 0.01$); $I^2 = 93\%$ | | | | |
| **Total (95% CI)** | | | **100.0%** | **0.93 [0.88; 0.98]** |
| **Prediction interval** | | | | **[0.77; 1.12]** |

Heterogeneity: $Tau^2 = 0.0069$; $Chi^2 = 163.02$, df = 13 ($P < 0.01$); $I^2 = 92\%$

Test for subgroup differences: $Chi^2 = 19.10$, df = 2 ($P < 0.01$)

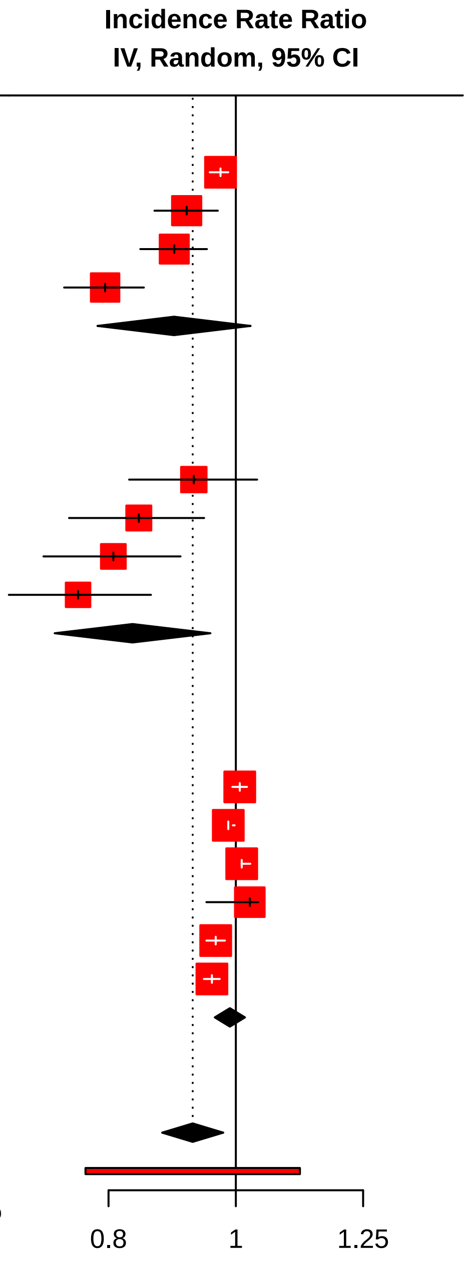
