## Supplemental Figure for "A global meta-analysis of effects of green infrastructure on COVID-19 infection and mortality rates": Figure S11.pdf

| Study or Subgroup | logIRR | SE | Weight | Incidence Rate Ratio IV, Random, 95% CI |
|---|---|---|---|---|
| economic = none | | | | |
| russette | −0.0862 | 0.0284 | 7.4% | 0.92 [0.87; 0.97] |
| russette | −0.1079 | 0.0298 | 7.3% | 0.90 [0.85; 0.95] |
| russette | −0.2291 | 0.0357 | 7.0% | 0.80 [0.74; 0.85] |
| Total (95% CI) | | | **21.6%** | **0.87 [0.72; 1.05]** |
| Heterogeneity: $Tau^2$ = 0.0047; $Chi^2$ = 10.66, df = 2 (P < 0.01); $I^2$ = 81% | | | | |
| economic = economic | | | | |
| klompmaker | −0.0269 | 0.0083 | 8.2% | 0.97 [0.96; 0.99] |
| sikarwar | −0.0737 | 0.0573 | 5.6% | 0.93 [0.83; 1.04] |
| sikarwar | −0.1701 | 0.0604 | 5.4% | 0.84 [0.75; 0.95] |
| sikarwar | −0.2147 | 0.0613 | 5.3% | 0.81 [0.71; 0.91] |
| sikarwar | −0.2765 | 0.0636 | 5.2% | 0.76 [0.67; 0.86] |
| yang | 0.0069 | 0.0065 | 8.2% | 1.01 [0.99; 1.02] |
| yang | −0.0132 | 0.0008 | 8.2% | 0.99 [0.99; 1.00] |
| yang | 0.0103 | 0.0039 | 8.2% | 1.01 [1.01; 1.03] |
| yang | 0.0245 | 0.0232 | 7.6% | 1.02 [0.95; 1.04] |
| yang | −0.0353 | 0.0083 | 8.2% | 0.97 [0.95; 0.98] |
| yang | −0.0419 | 0.0071 | 8.2% | 0.96 [0.95; 0.97] |
| Total (95% CI) | | | **78.4%** | **0.95 [0.89; 1.01]** |
| Heterogeneity: $Tau^2$ = 0.0056; $Chi^2$ = 109.46, df = 10 (P < 0.01); $I^2$ = 91% | | | | |
| **Total (95% CI)** | | | **100.0%** | **0.93 [0.88; 0.98]** |
| **Prediction interval** | | | | **[0.77; 1.12]** |

Heterogeneity: $Tau^2$ = 0.0069; $Chi^2$ = 163.02, df = 13 (P < 0.01); $I^2$ = 92%

Test for subgroup differences: $Chi^2$ = 2.60, df = 1 (P = 0.11)

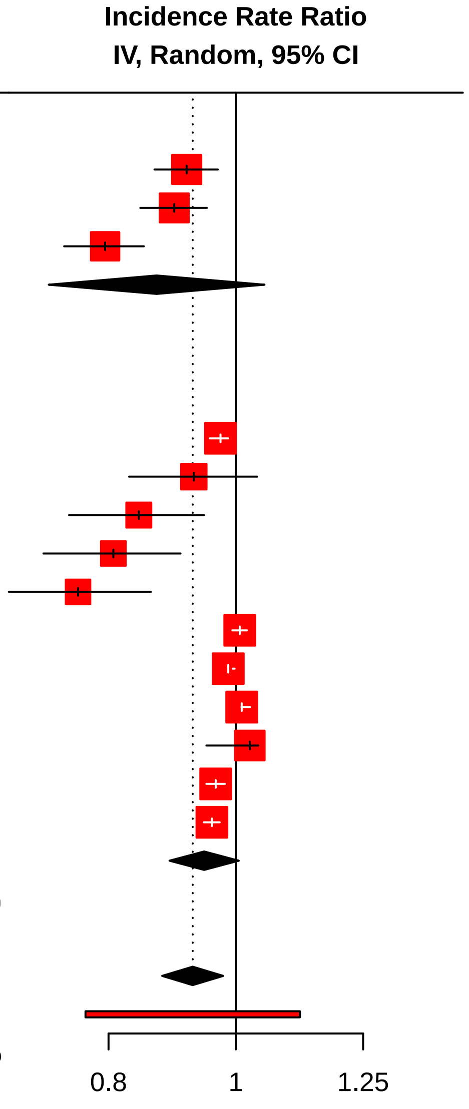
